# Added Value of Software-Assisted Analysis in FDG-PET for Neurodegenerative Disease Diagnosis: A Systematic Review and Meta-Analysis

**DOI:** 10.64898/2026.06.01.26354659

**Authors:** Hai-Jeon Yoon, Yeongjoo Lee, Ji-In Bang, Seo Young Kang, Ji-Young Kim, Miyoung Choi, Kyoungjune Pak

## Abstract

**Background:** In clinical practice, 18F-fluorodeoxyglucose positron emission tomography (FDG-PET) evaluation of neurodegenerative diseases relies primarily on visual interpretation, which is inherently subjective. Although current international guidelines recommend incorporating quantitative tools to support visual reading, the magnitude of the incremental diagnostic benefit and the clinical contexts in which it is most pronounced have not been formally synthesized in a systematic meta-analytic framework.

**Methods:** Following the Preferred Reporting Items for Systematic reviews and Meta-Analyses for Diagnostic Test Accuracy (PRISMA-DTA) guidelines, we searched PubMed, EMBASE, Cochrane Library, and KoreaMed from inception to August 2025 for studies comparing visual-only versus visual-plus-quantitative FDG-PET interpretation within identical patient cohorts. Pooled sensitivity and specificity were estimated using random-effects models. Relative diagnostic performance was summarized as odds ratios (ORs), obtained by exponentiation posterior contrasts between visual analysis combined with quantitative analysis, and visual analysis. Subgroup analyses were conducted based on the clinical experience of the readers.

**Results:** Ten studies met the inclusion criteria. In the overall analysis (k = 9), visual analysis alone yielded a pooled sensitivity of 0.85 (95% Confidence interval: 0.77–0.91) and specificity of 0.78 (0.63–0.88), versus a sensitivity of 0.87 (0.82–0.91) and specificity of 0.88 (0.74–0.95) for the combined approach. The most pronounced gain was observed in differentiating Alzheimer’s disease (AD) from healthy controls, with specificity increasing from 0.69 to 0.94 (Bayesian OR 4.29 (95% Credible interval: 1.87–10.84)). Quantitative augmentation conferred a larger sensitivity gain among beginner readers (increasing from 0.75 to 0.87; Bayesian OR 2.39 [1.33–4.32]) than among expert readers, narrowing the performance gap between experience levels.

**Conclusion:** Adding quantitative analysis to visual FDG-PET interpretation yields modest overall improvements in diagnostic accuracy, with the largest gains observed in distinguishing AD from cognitively normal individuals and among less experienced readers. These findings are consistent with current international guidelines that position quantitative assessment as a complementary aid to visual interpretation rather than a replacement, with particular utility for less experienced practitioners and for specific differential-diagnostic scenarios.

## INTRODUCTION

Brain 18F-fluorodeoxyglucose positron emission tomography (FDG-PET) is an established functional imaging biomarker for the differential diagnosis of neurodegenerative disorders^1^. By mapping regional cerebral glucose metabolism, FDG-PET captures synaptic dysfunction that often precedes detectable structural brain change, and characteristic hypometabolic patterns help discriminate Alzheimer’s disease (AD) from frontotemporal dementia (FTD) and dementia with Lewy bodies (DLB), as well as Parkinson’s disease (PD) from atypical parkinsonian syndromes such as multiple system atrophy (MSA), progressive supranuclear palsy (PSP), and corticobasal degeneration (CBD)^2, 3^. These properties are reflected in the appropriate use criteria and procedure guidelines of major international societies, which endorse FDG-PET as a supportive tool in the diagnostic work-up of patients with cognitive impairment and movement disorders of uncertain etiology^4, 5^. In current clinical practice, however, brain FDG-PET is interpreted primarily by visual inspection of cortical and subcortical metabolic patterns. Visual reading is inherently subjective and operator-dependent, and inter-reader agreement may be suboptimal for early-stage disease, atypical presentations, or mixed pathologies^6–8^.

To alleviate this subjectivity, quantitative approaches to FDG-PET reading—including z-score maps derived from comparison with age-matched normative databases, voxel-based statistical parametric analyses, and surface-projection–based regional metrics—have been developed to provide objective thresholds and to improve standardization across scanners and centers.^9–12^ These tools are widely used in research cohorts, and are increasingly being incorporated into routine clinical workflows as automated software packages become more widely available, a direction also endorsed by recent recommendations from the European Association of Nuclear Medicine (EANM) and the European Academy of Neurology (EAN), particularly to standardize reporting across centers and to support less experienced readers^2, 4, 12^. Previous studies suggest that augmenting visual interpretation with quantitative information can increase diagnostic confidence and reduce variability among readers, but reported effects on diagnostic accuracy have been inconsistent and often limited by small sample sizes and methodological heterogeneity.^12, 13^

Despite this growing clinical interest, evidence directly quantifying the incremental diagnostic value of combined visual-plus-quantitative interpretation over visual interpretation alone remains fragmented. The 2018 EANM–EAN consensus review, which surveyed the literature available up to 2015, identified only a small number of studies that performed a direct head-to-head comparison between visual and automated assessment in the same patient cohort, and acknowledged the need for procedural standardization.^12^ A more recent systematic review of FDG-PET in dementia subtype differentiation summarized overall diagnostic accuracy across dementia categories but did not specifically synthesize within-cohort comparisons of visual versus visual-plus-quantitative reading.^14^ Whether the benefit of adding quantitative analysis is uniform across diagnostic scenarios (e.g., AD/mild cognitive impairment (MCI) versus cognitively normal controls, AD/MCI versus other dementias, or PD versus atypical Parkinsonisms) and whether it differs by reader expertise have not been formally evaluated in a systematic meta-analytic framework.

To address these gaps, we conducted a systematic review and meta-analysis of studies that reported both visual-only and visual-plus-quantitative FDG-PET interpretations within identical patient cohorts (fully paired within-subject design). Using random-effects logistic-normal generalized linear mixed models for pooled estimates and Bayesian bivariate binomial-logit mixed-effects modeling for relative odds ratios, we compared pooled sensitivity and specificity between the two reading strategies in the overall population and in prespecified subgroups defined by diagnostic scenario and reader expertise. By restricting inclusion to paired head-to-head designs, this study aims to clarify the incremental diagnostic value of quantitative analysis as an adjunct to visual reading and to inform its optimal role in the clinical interpretation of brain FDG-PET in neurodegenerative disorders.

## MATERIALS AND METHODS

### Data Search and Study Selection

We conducted systematic reviews of EMBASE, PubMed, Cochrane, and KoreaMed databases from their inception through August 2025. The search strategy was structured using a combination of Medical Subject Headings (MeSH), Emtree terms, and natural language keywords to capture the following components: (1) population (P): neurodegenerative diseases, including Alzheimer’s, Parkinson’s, Huntington’s disease, multiple system atrophy, and other movement disorders; (2) intervention (I-1): 18F-FDG PET imaging, including terms for positron emission tomography and cerebral glucose metabolic rate; and (3) intervention (I-2): quantitative analysis methods such as statistical parametric mapping (SPM), stereotactic surface projection (SSP), radiomics, and artificial intelligence–based automated image analysis. The protocol was registered with the International Prospective Register of Systematic Reviews (PROSPERO; registration number: CRD420251112431).

### Inclusion and Exclusion Criteria

Our search was restricted to human subjects and research papers published in English. We specifically targeted studies assessing the role of quantitative FDG PET in the clinical evaluation of neurodegenerative conditions. To ensure the findings reflect real-world clinical practice, we included only those studies that compared visual-only interpretation versus visual-plus-quantitative interpretation (e.g., SPM, SSP, or AI-assisted tools) within the same patient cohort. The following were excluded from the final selection: (1) analysis methodology: studies that compared visual interpretation directly against quantitative analysis alone (excluding the hybrid approach used in clinical settings); (2) study type: review articles, conference abstracts, editorials, and animal studies; and (3) data integrity: duplicate publications and redundant reports from the same institution. If multiple reports from the same institution were found, only the most comprehensive or relevant study was included in each analysis.

### Review Process

Two researchers independently performed the initial search and screened titles and abstracts for eligibility. Following the initial screen, full-text articles were retrieved and reviewed to ensure they met the predefined inclusion criteria. Any discrepancies or disagreements between the researchers regarding study inclusion were resolved through consensus and discussion. Data extracted from each eligible study included publication year, country, study design, target disease(s), reference standard, sample size in each diagnostic group, characteristics and number of readers, quantitative method and software used, and the 2 × 2 contingency table for both the visual-only and visual-plus-quantitative interpretation arms.

### Quality Assessment

The methodological quality of each included study was assessed independently by two reviewers using the Quality Assessment of Diagnostic Accuracy Studies–2 (QUADAS-2) tool and its comparative extension, QUADAS-C, which is specifically designed for studies comparing two index tests within the same patients.^15, 16^ For each study, four QUADAS-2 domains (patient selection, index test, reference standard, and flow and timing) were rated for risk of bias and applicability concerns, and the corresponding QUADAS-C domains (patient selection comparison, index test 1 vs. index test 2 comparison, reference standard, and flow and timing comparison) were rated for risk of bias in the comparative assessment between visual-only and visual-plus-quantitative interpretation. Disagreements between the two reviewers were resolved by consensus, with arbitration by a third reviewer when necessary.

### Statistical Analysis

For diagnostic accuracy outcomes, pooled sensitivity and specificity were estimated separately for visual-only interpretation and for visual-plus-quantitative interpretation using random-effects logistic-normal generalized linear mixed models (GLMMs) with logit transformation, implemented with the metaprop function of the meta package. Direct, head-to-head comparisons between visual-only and visual-plus-quantitative interpretation within the same studies were performed using a Bayesian bivariate binomial-logit generalized linear mixed-effects model fitted with the brms package. In this model, the numbers of true positives (TP) and true negatives (TN) were jointly modeled as binomial outcomes with logit link, with arm (visual-only vs. visual-plus-quantitative) as a fixed effect and a study-level random intercept that allowed correlation between the sensitivity and specificity dimensions, thereby explicitly accounting for the paired within-study structure. Weakly informative priors were specified for fixed effects, random-effect standard deviations, and the between-response correlation. Bayesian models were estimated using four Markov chains, each consisting of 4,000 iterations with a 1,000-iteration warm-up period, resulting in a total of 12,000 post-warmup samples. Relative diagnostic performance was summarized as odds ratios (ORs), obtained by exponentiation of posterior contrasts between visual-plus-quantitative and visual-only interpretation on the logit scale. As a secondary analysis, relative sensitivity and specificity were additionally evaluated using a frequentist random-effects meta-regression of logit-transformed proportions with study-level paired random effects, and effect estimates were reported as ORs. Bayesian estimates are reported with 95% credible intervals, whereas frequentist estimates are reported with 95% confidence intervals. Subgroup analyses were conducted based on 1) neurodegenerative disease, and 2) the clinical experience of the readers. All statistical analyses were performed using R version 4.5.2 (R Foundation for Statistical Computing, Vienna, Austria).

## RESULTS

### Study Selection and Study Characteristics

The systematic search across EMBASE, PubMed, Cochrane, and KoreaMed databases identified a total of 2,483 records. After removing 616 duplicate records, 1,295 records were excluded by title, and 572 records were screened by abstract. After excluding a further 512 records, 60 full-text reports were assessed for eligibility. Finally, 10 studies meeting the inclusion and exclusion criteria were included in the meta-analysis (Figure 1)^17–26^. Studies included in the meta-analysis are summarized in Table 1. The number of readers in each study varied from 1 to 6. The diagnostic groups evaluated primarily included patients with AD, MCI, and PD. Differential diagnosis groups included healthy controls (HC), patients with cerebrovascular disease, FTD, DLB, MSA, PSP, and CBD.

**Figure 1.**
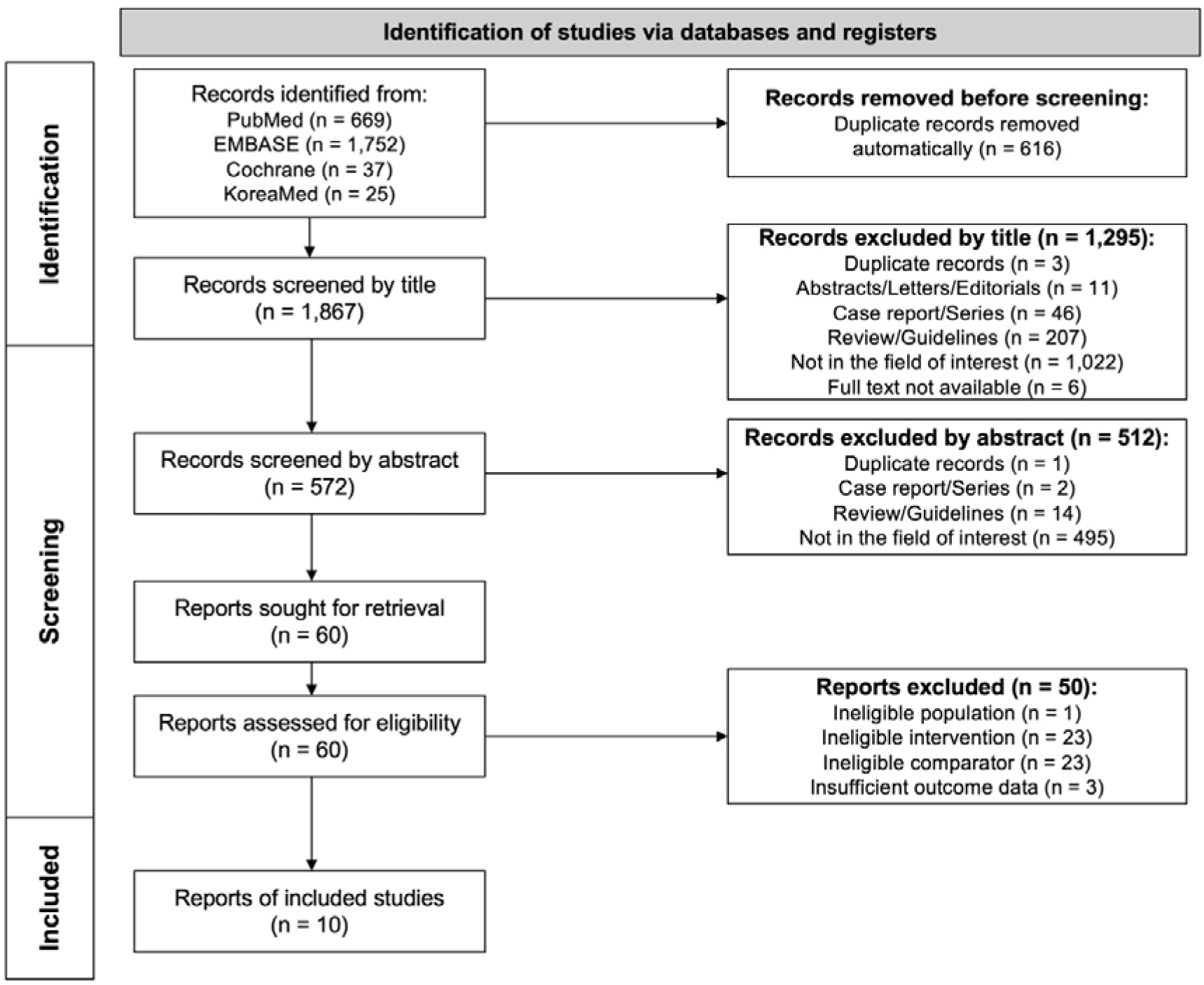
PRISMA flow diagram depicting the study selection process.

**Table 1.**
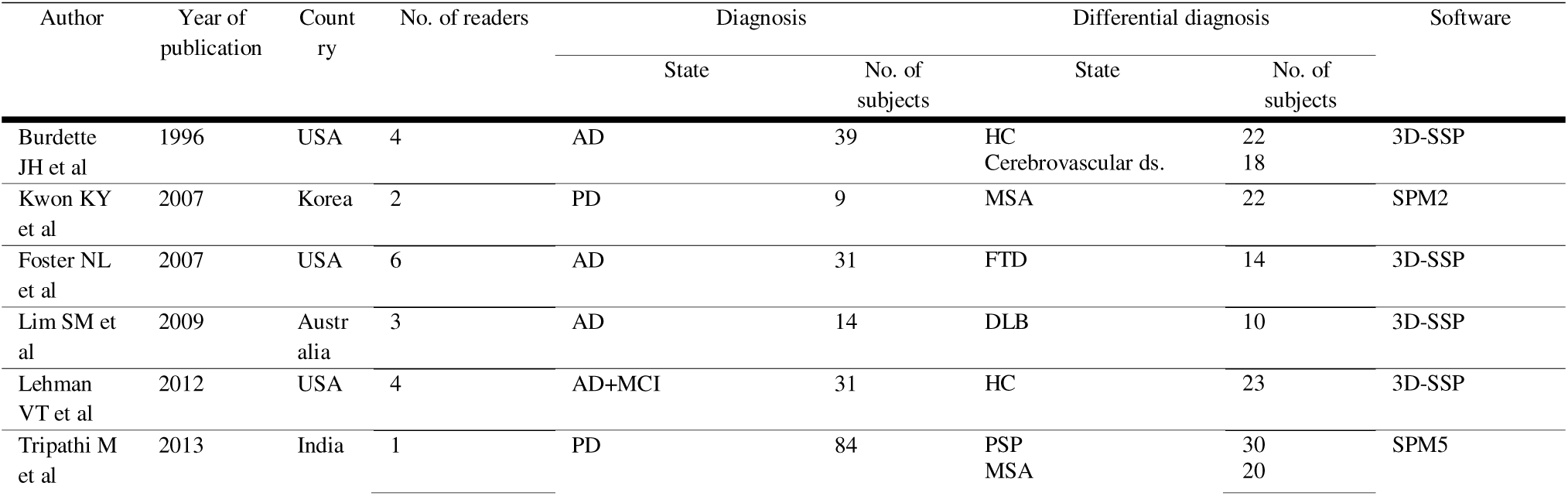
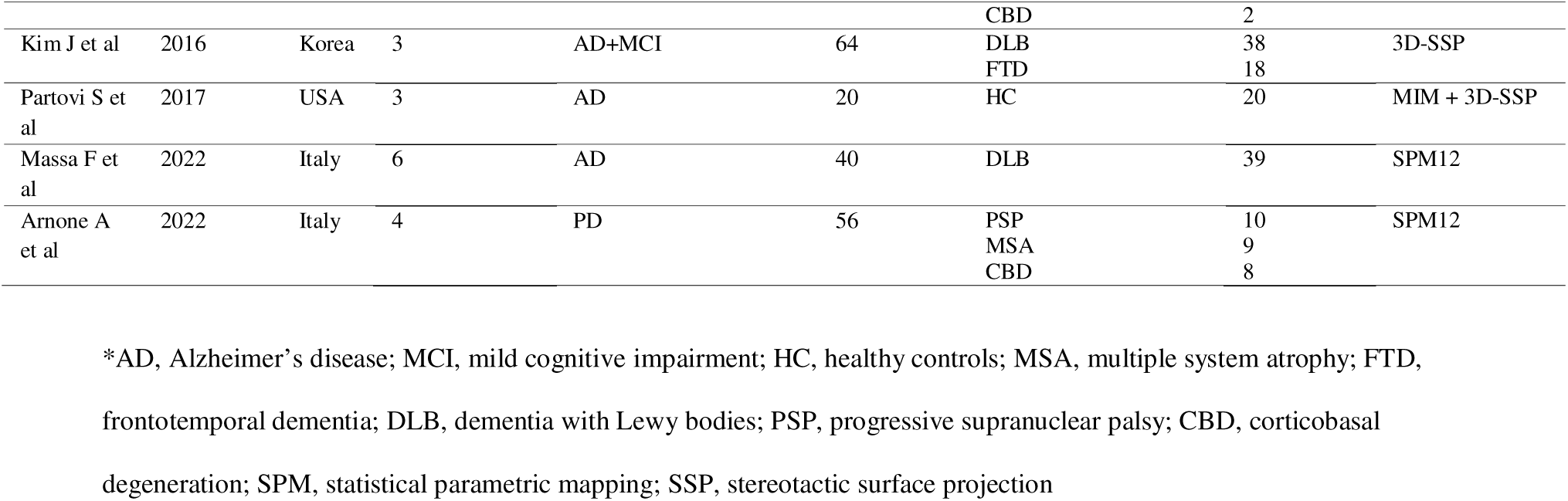
Characteristics of the included studies.

### Quality Assessment

The results of the methodological quality assessment using QUADAS-2 and QUADAS-C are summarized in Supplementary figure 1. In the QUADAS-2 assessment, seven of the ten studies were rated as having a high risk of bias in the patient selection domain, reflecting a case-control design or the use of pre-selected diagnostic groups that did not represent a consecutive clinical population^17, 19–24^. In the reference standard domain, one study was judged to carry an applicability concern because the clinical reference diagnosis was partly informed by the qualitative FDG-PET findings, raising the possibility of incorporation bias^26^. In the QUADAS-C assessment of the comparison between visual-only and visual-plus-quantitative interpretation, the index-test comparison was rated as having a high risk of bias in two studies, reflecting partial non-independence in studies where readers performed the two reading strategies in a stepwise manner.

### Diagnostic performance of visual-only vs visual-plus-quantitative interpretation

The diagnostic performance of visual-only interpretation versus visual-plus-quantitative interpretation was calculated. A total of nine studies included in the pooled analysis, as one study by Foster et al. was excluded from the primary pooled analysis owing to patient-cohort overlap with another included study by Burdette et al^17–19, 21–24, 26, 27^. Relative diagnostic performance was further assessed using both frequentist and Bayesian ORs, where values greater than 1.0 indicate improved performance with the addition of quantitative analysis. In the pooled analysis of 9 studies, visual-only interpretation demonstrated a sensitivity of 0.85 (95% confidence interval [CI]: 0.77–0.91) and a specificity of 0.78 (0.63–0.88). Visual-plus-quantitative interpretation showed a sensitivity of 0.87 (0.82–0.91) and a specificity of 0.88 (0.74–0.95). The corresponding frequentist OR was 1.38 (0.89–2.14) for sensitivity, and 1.66 (0.95–2.89) for specificity, while Bayesian OR was 1.29 (95% credible interval [CrI]: 0.86–1.94) for sensitivity and 1.64 (1.07–2.52) for specificity (Figure 2).

**Figure 2.**
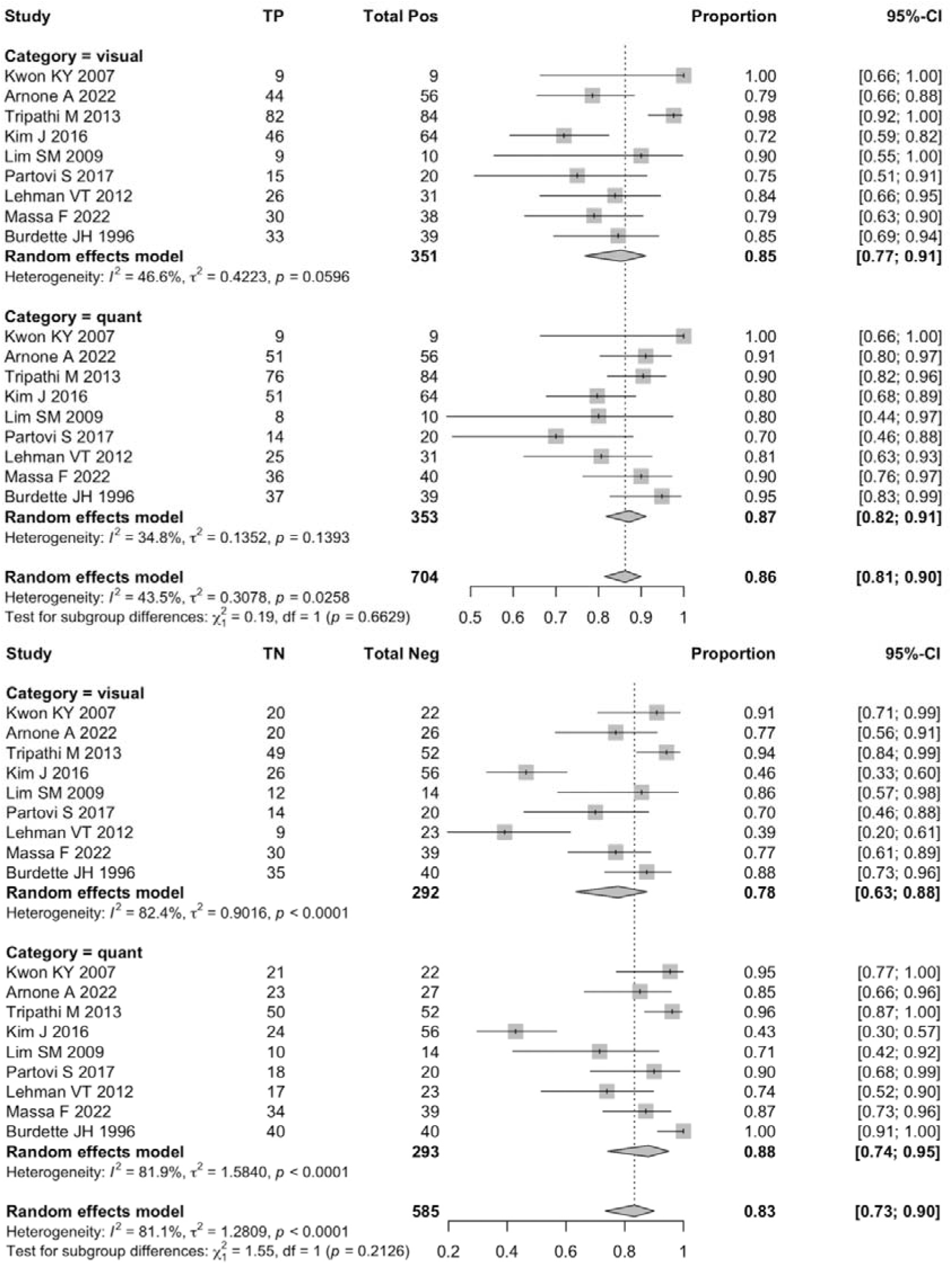
Forest plots comparing the diagnostic performance between visual-only interpretation (visual) versus visual-plus-quantitative interpretation (quant) in the overall pooled analysis (k = 9). Top: pooled sensitivity; bottom: pooled specificity. Results are shown for individual studies and as random-effects pooled estimates.

### Subgroup Analysis of Alzheimer’s Disease

In the differentiation of AD from HC,^17–19^ visual interpretation resulted in a sensitivity of 0.82 (0.73–0.89) and a specificity of 0.69 (0.41–0.88). Visual-plus-quantitative interpretation improved these values to a sensitivity of 0.85 (0.69–0.93) and a specificity of 0.94 (0.64–0.99). The corresponding frequentist OR was 1.04 (0.46–2.35) for sensitivity, and 4.39 (1.69–11.44) for specificity. The Bayesian OR was 1.16 (0.55–2.48) for sensitivity and 4.29 (1.87–10.84) for specificity. When comparing AD against non-AD dementias^20–23^, visual interpretation yielded a sensitivity of 0.85 (0.69–0.93) and a specificity of 0.66 (0.48–0.81). Visual-plus-quantitative interpretation showed a sensitivity of 0.88 (0.77–0.94) and a specificity of 0.70 (0.48–0.85). The corresponding frequentist OR was 1.56 (0.82–2.99) for sensitivity, and 1.08 (0.62–1.89) for specificity. The Bayesian OR for this comparison was 1.51 (0.81–2.81) for sensitivity and 1.08 (0.63–1.84) for specificity.

### Subgroup Analysis of Parkinson’s Disease

In a pooled analysis of studies with PD,^24, 26, 27^ visual interpretation showed a sensitivity of 0.94 (0.73–0.99) and a specificity of 0.89 (0.78–0.95), while visual-plus-quantitative interpretation yielded a sensitivity of 0.91 (0.86–0.95) and a specificity of 0.93 (0.85–0.97). The corresponding frequentist OR was 1.01 (0.17–5.94) for sensitivity, and 1.70 (0.62–4.68) for specificity. The Bayesian OR for this group was 1.08 (0.51–2.30) for sensitivity and 1.58 (0.63–4.05) for specificity.

### Subgroup Analysis by Reader Experience

Subgroup analyses were conducted based on the clinical experience of the readers^18, 22, 26^. For expert readers, visual-only interpretation yielded a sensitivity of 0.82 (0.65–0.92) and a specificity of 0.65 (0.56–0.74). Visual-plus-quantitative interpretation increased sensitivity to 0.87 (0.73–0.95) and specificity to 0.75 (0.50–0.91), yielding a frequentist OR of 1.61 (0.86–3.02) for sensitivity and 1.49 (0.39–5.64) for specificity, and a Bayesian OR of 1.58 (0.87–2.92) for sensitivity and 1.05 (0.60–1.84) for specificity (Figure 3). The benefit of quantitative analysis was most pronounced among beginner readers. In this group, visual-only interpretation showed a sensitivity of 0.75 (0.62–0.84) and a notably low specificity of 0.41 (0.16–0.72). Visual-plus-quantitative interpretation improved sensitivity to 0.87 (0.81–0.92) and specificity to 0.60 (0.28–0.85). This resulted in a frequentist OR of 2.58 (1.40–4.74) for sensitivity and 1.96 (0.83–4.61) for specificity, and a Bayesian OR of 2.39 (1.33–4.32) for sensitivity and 1.81 (0.99–3.33) for specificity (Figure 4).

**Figure 3.**
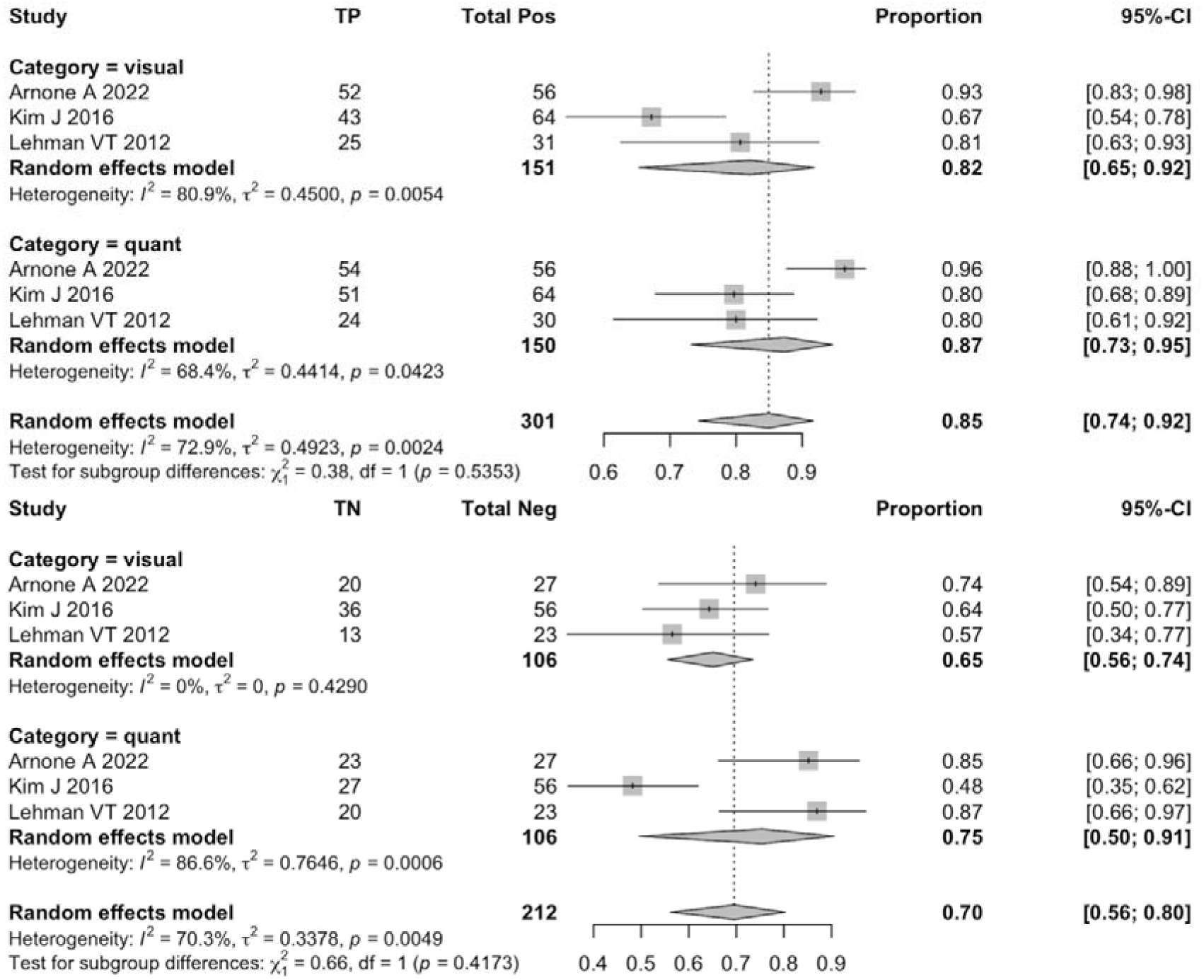
Forest plots comparing the diagnostic performance between visual-only interpretation versus visual-plus-quantitative interpretation in the expert-reader subgroup (k = 3). Top: pooled sensitivity; bottom: pooled specificity.

**Figure 4.**
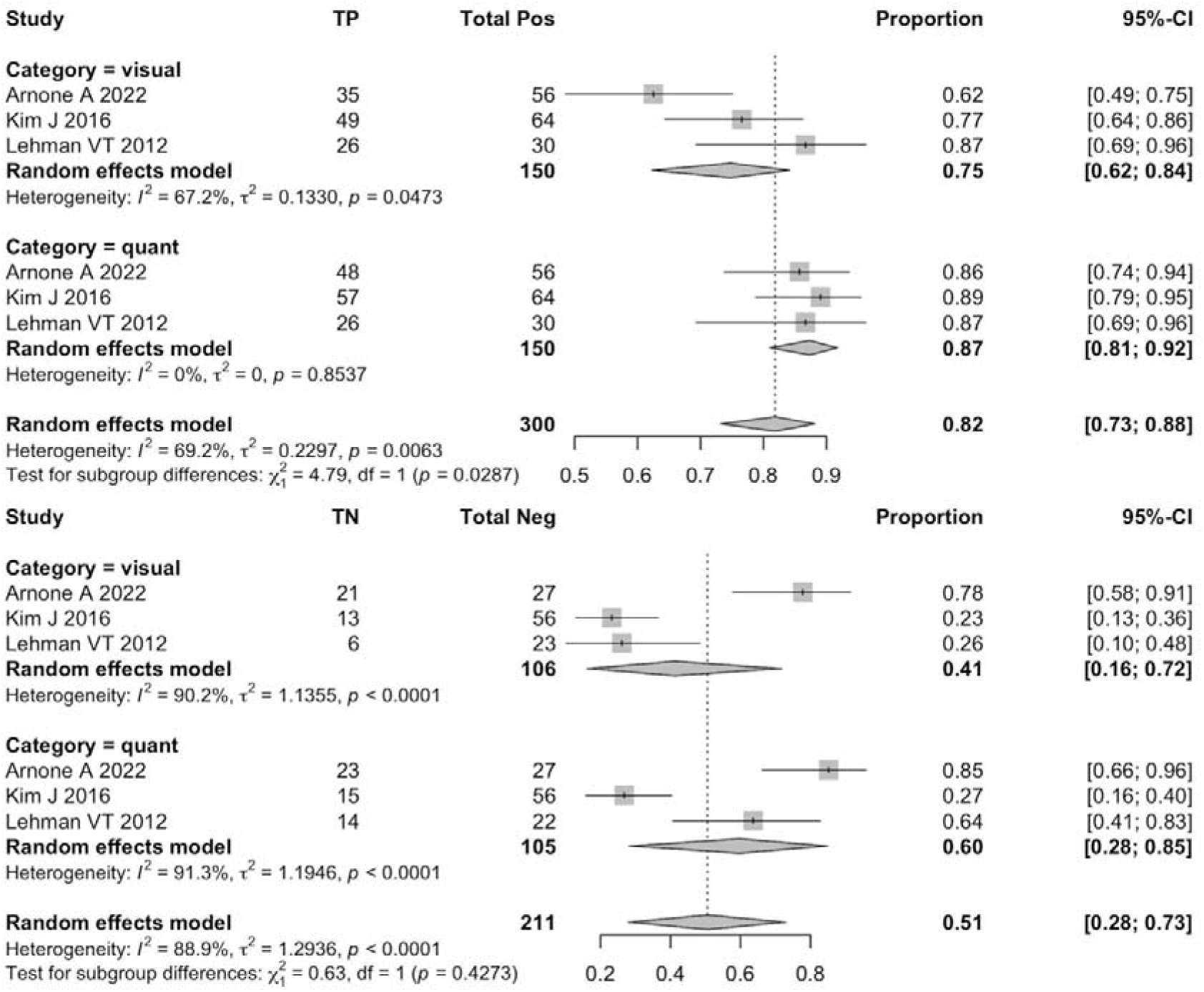
Forest plots comparing the diagnostic performance between visual-only interpretation versus visual-plus-quantitative interpretation in the beginner-reader subgroup (k = 3). Top: pooled sensitivity; bottom: pooled specificity.

## DISCUSSION

In this systematic review and meta-analysis of 10 head-to-head comparison studies between visual-only and visual-plus-quantitative FDG-PET interpretation, we found that the addition of quantitative analysis produced a modest but clinically meaningful improvement in diagnostic accuracy. In the overall analysis, the gain was driven primarily by specificity, which increased from 0.78 to 0.88 (Bayesian OR 1.64, 95% CrI 1.07–2.52), while sensitivity remained essentially unchanged. The magnitude and clinical relevance of this benefit, however, varied substantially across diagnostic scenarios and reader experience levels, indicating that the incremental value of quantitative analysis is context-dependent rather than uniform.

Our findings are consistent with and extend the conclusions of the 2018 EANM–EAN consensus, which observed that automated assessment generally improves specificity and marginally improves overall accuracy, while sensitivity remains comparable to visual reading.^12^. However, the 2018 EANM–EAN consensus was based on only three studies that performed a direct head-to-head comparison and therefore could not provide pooled effect estimates or stratified analyses. By restricting inclusion to paired within-subject designs and applying random-effects logistic-normal GLMMs together with a Bayesian bivariate binomial-logit mixed-effects model, the present meta-analysis provides quantitative effect estimates with credible intervals, allowing the magnitude and uncertainty of the incremental benefit to be characterized formally for the first time. Furthermore, a recent systematic review of FDG-PET for dementia subtype differentiation reported high overall diagnostic accuracy of FDG-PET across dementia categories but did not separately synthesize the within-cohort comparison between reading strategies^14^. Our study complements this work by focusing specifically on the comparative question of whether quantitative augmentation adds incremental value over visual reading alone.

In the differentiation of AD from cognitively normal individuals, the pooled analysis included three studies^17–19^, all of which employed a paired within-subject design comparing visual-only and visual-plus-quantitative interpretation on the same images. The most pronounced gain was observed in specificity, which increased from 0.69 to 0.94 (Bayesian OR 4.29, 95% CrI 1.87–10.84), suggesting that objective comparison against normative reference data through 3D-SSP projection or z-score mapping effectively offsets reader uncertainty in distinguishing the subtle hypometabolic patterns of AD from age-related normal variation, which is particularly meaningful given the substantial clinical implications of false-positive AD diagnoses in cognitively normal individuals for diagnostic work-up and patient counseling^8^.

The pooled analysis for differentiation of AD from non-AD dementias included four studies^20–23^. In the pooled analysis, both sensitivity (increasing from 0.85 to 0.88; Bayesian OR 1.51, 95% CrI 0.81–2.81) and specificity (increasing from 0.66 to 0.70; Bayesian OR 1.08, 95% CrI 0.63–1.84) showed only small and statistically non-significant gains, likely because the characteristic hypometabolic patterns of non-AD dementias such as FTD and DLB are sufficiently distinctive on visual reading by experienced readers, leaving limited incremental room for quantitative augmentation to contribute^6^. However, these studies evaluated heterogeneous comparator conditions using a wide range of quantitative methods, resulting in substantial clinical and methodological heterogeneity.

The pooled analysis of Parkinsonian syndromes included three studies^24–26^. In the pooled analysis, both arms achieved specificity exceeding 89% (0.89 for visual-only, 0.93 for visual-plus-quantitative), and the incremental gain from quantitative augmentation was modest (Bayesian OR 1.58 for specificity, 1.08 for sensitivity), consistent with the interpretation that the characteristic metabolic patterns of parkinsonian syndromes—such as putaminal hypometabolism in MSA-P or cerebellar involvement in MSA-C—are visually recognizable with reasonable confidence even without quantitative support. Importantly, the study by Arnone et al. carried a high applicability concern for the reference standard because the clinical diagnosis was supported by FDG-PET results.^26^

The subgroup analysis by reader experience included three studies^18, 22, 26^. In the pooled analysis, the addition of quantitative analysis yielded only a modest improvement in sensitivity among expert readers (increasing from 0.82 to 0.87; Bayesian OR 1.58, 95% CrI 0.87–2.92), whereas among beginner readers the gain was statistically significant and substantially larger, with sensitivity increasing from 0.75 to 0.87 (Bayesian OR 2.39, 95% CrI 1.33–4.32) and specificity rising from 0.41 to 0.60. This pattern empirically demonstrates that quantitative tools deliver the greatest added value where reader expertise is most variable, narrowing the diagnostic performance gap across experience levels and providing concrete support for the EANM–EAN recommendation in this context^2, 4, 12^. However, this subgroup is based on only three studies, which limits statistical power, and the definitions of “expert” versus “beginner” were not standardized across studies, so these estimates should be interpreted with caution.

Although the studies by Burdette et al. and Foster et al. were identified as having been conducted at the same institution, the study by Foster et al. was selectively excluded from the pooled primary analysis to avoid double-counting of overlapping AD patient cohorts^17, 20^. For the subsequent subgroup analyses, however, both reports were included independently as they evaluated distinct clinical comparisons; Burdette et al. focused on the differentiation of AD from HC and cerebrovascular disease, whereas Foster et al. addressed the differentiation of AD from FTD. This approach maximizes the use of the available within-cohort comparative information while preserving the methodological integrity of the primary effect estimate.

Taken together, our findings support a context-aware use of quantitative analysis as a complement, rather than a substitute, for visual interpretation of brain FDG-PET in neurodegenerative disorders. In scenarios where visual reading is already highly accurate, such as PD versus atypical parkinsonian syndromes in our analysis, where both arms exceeded 89% specificity, the marginal benefit of quantitative augmentation may be limited. In contrast, in scenarios with a high prior probability of borderline or atypical hypometabolic patterns, or when scans are interpreted by less experienced readers, quantitative information appears to provide a measurable diagnostic gain.

Several limitations should be acknowledged. First, the number of eligible head-to-head studies remained limited (k = 10), reflecting the relative scarcity of paired within-subject designs in this field; this restricted the statistical power for some subgroup comparisons, particularly the reader-experience subgroup based on three studies. Second, the included studies used a range of quantitative methods and normative databases, which may contribute to between-study heterogeneity even within the comparative effect estimates. Third, several studies adopted case-control designs; although such designs can inflate both sensitivity and specificity through spectrum bias, their reliance on clearly defined control groups, whether healthy controls or pre-confirmed distinct disease groups such as non-AD dementias or atypical parkinsonian syndromes, bears most directly on specificity in this setting. Fourth, our search was restricted to English-language publications, which may have led to the omission of relevant non-English studies. Finally, inter-reader agreement could not be synthesized quantitatively owing to inconsistent reporting across studies; this represents an important direction for future head-to-head research.

## CONCLUSION

Adding quantitative analysis to visual interpretation of brain FDG-PET in patients with suspected neurodegenerative disorders provides a modest but clinically meaningful improvement in diagnostic accuracy, driven primarily by gains in specificity. The benefit is heterogeneous across clinical contexts, with the largest gains in differentiating AD from cognitively normal individuals and among less experienced readers. These findings support the integration of quantitative tools into routine FDG-PET reporting workflows as a complementary aid to visual reading rather than a replacement, with particular utility for specific differential-diagnostic scenarios and for settings with variable reader expertise.

## Data Availability

All data produced in the present study are available upon reasonable request to the authors

